# Strategy for a rapid screening and surveillance of SARS-CoV-2 variants by real time RT-PCR: a key tool that allowed control and delay in Delta spread in Cordoba, Argentina

**DOI:** 10.1101/2021.11.16.21266265

**Authors:** Gonzalo M. Castro, Paola Sicilia, María Laura Bolzon, Laura López, María Gabriela Barbás, María Belén Pisano, Viviana E. Ré

**Author notes:** Corresponding author: María Belén Pisano, Enfermera Gordillo Gómez s/n, Ciudad Universitaria, Córdoba, Argentina. CP: 5016.

## Abstract

**Background:** SARS-CoV-2 variants of concern (VOC) and interest (VOI) present mutations in reference to the original virus, being more transmissible. We implemented a rapid strategy for the screening of SARS-CoV-2 VOC/VOIs using real time RT-PCR and performed monitoring and surveillance of the variants in our region.

**Methods:** consecutive real-time RT-PCRs for detection of the relevant mutations/deletions present in the Spike protein in VOC/VOIs (TaqMan™ SARS-CoV-2 Mutation Panel, Applied Biosystems) were implemented. An algorithm was established and 3941 SARS-CoV-2 RNA samples (Cts<30) obtained from oropharyngeal swabs from infected individuals in Córdoba, Argentina, between January and October 2021, were analyzed.

**Results:** the strategy of choice included a first screening of 3 mutations (N501Y, E484K, L452R) followed by the detection of other mutations/deletions based on the results. The analyses of the samples showed introductions of VOCs Alpha and Gamma in February and March 2021, respectively. Since then, Alpha presented a low to moderate circulation (1.7% of the SARS-CoV-2 currently detected). Gamma showed an exponential increase, with a peak of detection in July (72%), until reaching a current frequency of 41.1%. VOC Delta was first detected in July in travellers and currently represents 35% of detections in the community. VOI Lambda presented a gradual increase, showing a current frequency of 29%.

**Conclusions:** we report a useful tool for VOC/VOI detection, innovative for Argentina, capable to quickly and cost-effectively monitor currently recognized variants. It was key in the early detection of Delta, being able to implement measures to delay its dissemination.

## Introduction

Numerous severe acute respiratory syndrome coronavirus 2 (SARS-CoV-2) variants have already been documented globally during the COVID-19 pandemic. These viruses present one or more mutations in reference to the original virus, first isolated in Wuhan in 2019 [Janik et al. 2021] (consensus sequence WIV04) [GISAID]. Most nucleotide changes have little to no impact on the virus’ properties; however, there are mutations that produce phenotypic changes, which may affect virus transmissibility, severity, response to the vaccine or diagnostic tools [WHO 2021]. According to the virus’s features given by the mutations, the World Health Organization (WHO) and the Centers for Disease Control and Prevention (CDC) have classified some SARS-CoV-2 variants into variants of concern (VOC) and variants of interest (VOI) [WHO 2021, CDC 2021]. Thus, the first VOC described was the Alpha variant (lineage B.1.1.7), detected firstly in United Kingdom on September 2020, which rapidly expanded and become predominant in many countries, being the main variations the HV 69-70 deletion and N501Y mutation in the Spike protein [Janik 2021, Ong et al. 2021] (Figure 1A). After that, VOCs Beta (lineage B.1.351) and Gamma (lineage P.1) were reported, first isolated in South Africa and Brazil, respectively, presenting high transmissibility rates too, and with the common mutation N501Y (Figure 1A). Delta variant is the last VOC described up to date, detected for the first time in India in October 2020 and declared VOC on May 2021, which has displaced the rest of the VOCs in many parts of the world, with the presence of the main mutations L452R and P681R among others [Boehm et al. 2021, Outbreak info 2021] (Figure 1A). Over the last few months, several variants were classified as of interest (VOI), but later demonstrated to no longer pose a major added risk to global public health compared to other circulating SARS-CoV-2 variants, so they were re-classified as variants under monitoring (VUM) or even removed from the VOI/VUM list (OMS, 2021). At present, there are 2 VOIs: Lambda and Mu (OMS 2021). Lambda (lineage C.37), first detected in Peru on December 2020, was declared VOI on June 2021, and presents the characteristic mutation L452Q. Mu (lineage B.1.621), first isolated in Colombia and declared VOI at the end of August 2021, presents the main mutations N501Y, E484K and P681H (OMS 2021, Outbrek info). Former VOIs Zeta and Epsilon were declared of interest between March and July 2021 and present the mutations E484K and L452R, respectively.

**Figure 1.**
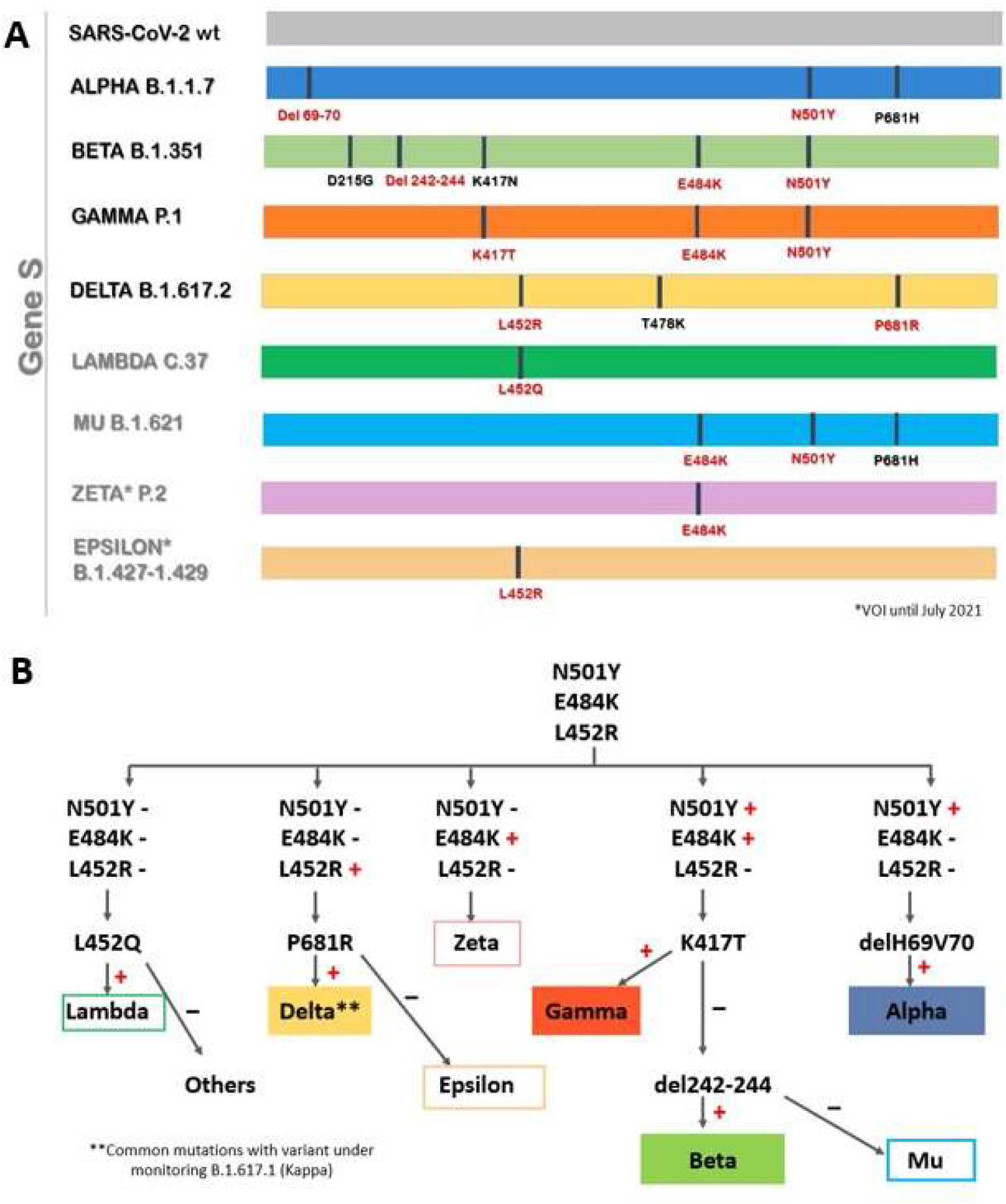
Some of the relevant mutations present in SARS-CoV-2 variants of concern (VOC) and variants of interest (VOI) (A), and VOC/VOI detection strategy implemented in this study, using consecutive real time RT-PCRs for relevant mutation screening (B).

Due to the increased transmissibility, possible increased virulence or changes in clinical disease presentation, and immune escape [Boehm et al. 2021], VOC/VOIs have the highest priority for surveillance, either to describe its circulation, map their spread, and to detect the introduction of new variants in a region. Currently, this surveillance is based on whole genome sequencing (WGS), an accurate method that generates detailed information, considered the gold standard technique to detect VOC/VOIs [Ong et al. 2021, Lind et al. 2021]. However, it is time-consuming, expensive, and requires trained staff and specific equipment, so it is not possible to apply it massively or to obtain results in a short-period time [Ong et al. 2021].

Since March 2020, in Argentina, and particularly in Córdoba Province, molecular surveillance of SARS-CoV-2 began with WGS [Proyecto PAIS]. As a result of increased concern in public health due to the emergent variants, in the last months there has been a rapidly growing demand for WGS. In February 2021, the strategy of partial sequencing of the S gene was implemented for the search of VOC/VOIs in our region, and VOCs Alpha and Gamma, as well as former VOI Zeta were detected for the first time in our province in March 2021 [Reporte N°18 proyecto PAIS]. From the Epidemiology Area of the Government of Córdoba Province, blocking actions have been carried out by tracking the index cases and close contacts for their isolation and follow-up, which were incremented since VOCs entered into our region. However, since WGS and partial sequencing of the S gene are time-consuming methods, they do not allow the rapid identification of VOCs, delaying their spread control. Added to this, the demand for variant typing increased as Delta was declared a VOC and began to spread around the world (WHO, 2021). Based on these facts, it was necessary to provide a rapid typing response, from the laboratory, in order to take measures at the public health level to contain the spread of Delta.

In this context, we developed a strategy using real time reverse transcriptase polymerase chain reactions (RT-PCR) for detection of VOC/VOI relevant mutations (using specific probes). In this article, we describe our experience and show the results obtained during the molecular surveillance carried out with this methodology, which was key to control and delay of VOC Delta dissemination in the Central region of Argentina.

## Methods

### Samples

A total of 3941 positive SARS-CoV-2 RNAs with Cts<30, obtained from oropharyngeal swab samples from individuals from the province of Córdoba, Argentina, between 1^st^ January and 31^st^ October 2021 were selected to screen VOC and VOI mutations by real time RT-PCR. The samples had originally been extracted with MegaBio plus Virus RNA Purification Kit II (BioFlux) on the GenePure Pro Nucleic Acid Purification System NPA-32P and amplified by real time RT-PCR using DisCoVery SARS-CoV-2 Nucleic Acid Detection Kit. Samples were randomly selected from all the province and from all the months studied in order to monitor VOC/VOI circulation in the community. Six hundred and seventy-seven samples corresponded to travellers who arrived to Córdoba and to samples obtained from the study of outbreaks associated with VOC Delta, analysed with the aim to carry out a rapid surveillance of VOC/VOIs which allows decision-making regarding sanitary measures.

### Detection of VOCs by real time PCR

TaqMan™ SARS-CoV-2 Mutation Panel (Applied Biosystems) was used for detection of the following relevant mutations/deletions present in VOC/VOIs: HV 69-70 del, N501Y, E484K, L452R, K417T, P681R, 242-244 del, L452Q. Briefly, 7μL of RNA were added to 8 μL of a mixture containing TaqPath™ 1-Step RT-qPCR Master Mix, CG (4X), TaqMan™ SARS-CoV-2 Mutation Panel Assay (40X) and nuclease-free water.

A strategy for detection of VOC/VOIs was implemented based on the different mutations found for each of the variants. Former VOIs Zeta and Epsilon were also included only in the samples from January to July, when those variants were classified as VOIs. The following samples were used as reference (obtained in collaboration with PAIS Project) (Proyecto PAIS 2021): EPI_ISL_2271687 to EPI_ISL_2271690 (VOC Gamma), EPI_ISL_2007514 to EPI_ISL_2007516 (VOC Alpha), EPI_ISL_3230016 to EPI_ISL_3230018 (VOC Delta), EPI_ISL_3183944 (former VOI Epsilon), EPI_ISL_3183946 (VOI Lambda), EPI_ISL_3183947 (former VOI Zeta), EPI_ISL_6032791 (VOI Mu).

## Results

We implemented a strategy for detection of SARS-CoV-2 VOCs (Alpha, Beta, Gamma, Delta) and VOIs [Lambda and Mu; additionally, Zeta and Epsilon (VOIs until July 2021)] by real time RT-PCR looking for some characteristic mutations (Figure 1B). A first screening for N501Y, E484K and L452R mutations was carried out (Figure 1B). Based on the results, the following algorithm was continued:

a. N501Y(+), E484K(+), L452R(-): detection of K417T was performed; a positive result was indicative of VOC Gamma (N501Y, E484K, K417T). A negative result was subjected to detection of 242-244del: if the deletion was present, VOC Beta would be the infecting variant (N501Y, E484K, 242-244del). A negative result for K417T and 242-244del, in the presence of mutations N501Y and E484K, was indicative of VOI Mu.
b. N501Y(+), E484K(-), L452R(-): deletion 69-70 was investigated; a positive result was indicative of VOC Alpha.
c. N501Y(-), E484K(+), L452R(-): the presence of only the E484K mutation would rule out the presence of VOCs, and would be indicative of former VOI Zeta.
d. N501Y(-), E484K(-), L452R(+): detections of P681R was performed; a positive result was indicative of possible VOC Delta. A negative result for P681R was indicative of former VOI Epsilon.
e. N501Y(-), E484K(-), L452R (-): detection of L452Q was performed; if this mutation was present, VOI Lambda would be the variant.

Once this strategy was implemented, the processing of the samples began, and a total of 3941 samples was tested. Figure 2 shows changes in VOC/VOIs distribution within SARS-CoV-2 infections among the community in Córdoba during the studied period. VOCs Alpha and Gamma were first detected in our region in the months of February and March, respectively. From that moment, they began to circulate in the population. VOC Alpha was detected in all months, reaching a peak of 12% in May 2021. Then, it was detected in a less frequency, being the 1.7% of the samples tested from October 2021 (Figure 2). Since its first detection in March 2021, VOC Gamma presented an exponential rice, and became predominant in the following months, reaching a peak of detection on July 2021 (72%) (Figure 2). From that moment, this VOC started to decrease, and it is currently detected in the 41.1% of the samples of the community. VOC Beta was not detected.

**Figure 2.**
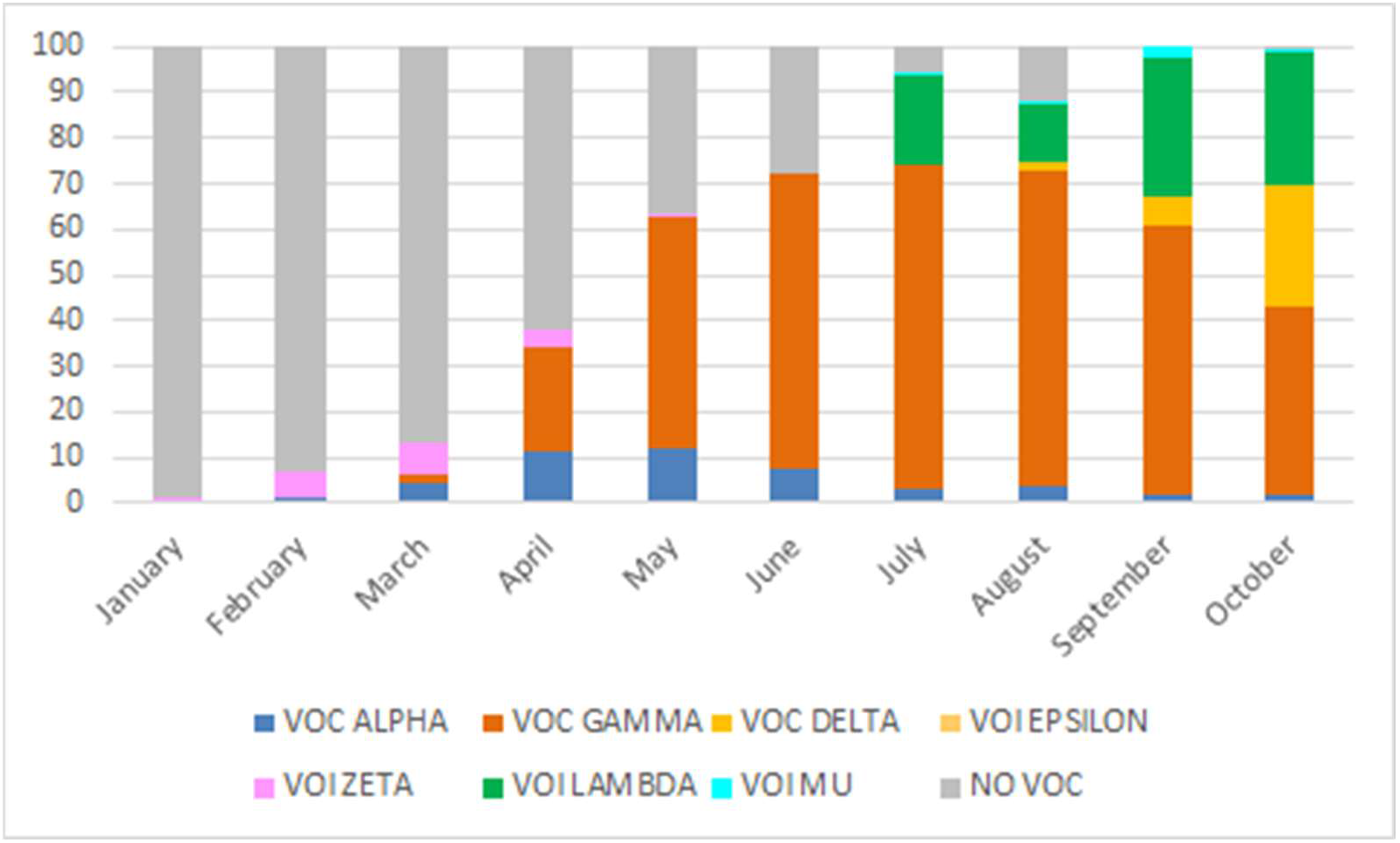
Distribution of SARS-CoV-2 variants of concern (VOC) and interest (VOI) detected in the community from Córdoba Province, Argentina, by real time RT-PCR, January to October 2021.

VOC Delta was first detected in a traveller from Peru on July 2021. On August 2021 it was identified in samples from the community in a frequency of 1.8%, increasing gradually up to 35% detected nowadays (Figure 2).

Regarding VOIs, Zeta circulated from January to May 2021 in a low frequency (1.1% to 0.7%, with a peak in May 2021 of 6.9%). Epsilon was only detected in August (0.2%). VOIs Lambda and Mu started to be screened on July 2021. Lambda was detected in the 19.2% of the samples that month, and it gradually increased its frequency until reaching 29% on October 2021. VOI Mu was detected in a low frequency (less than 3%).

On July 2021, the study of variants in travellers and their close contacts was intensified in our province, due to the opening of the country’s borders and the beginning of travels, added to the spread of VOC Delta in the world some weeks before. Figure 3 shows the results obtained from the typing of the samples in this group and in those samples obtained during the study of outbreaks associated to Delta (that began to be registered since this VOC entered into our province) (July to October 2021).

**Figure 3.**
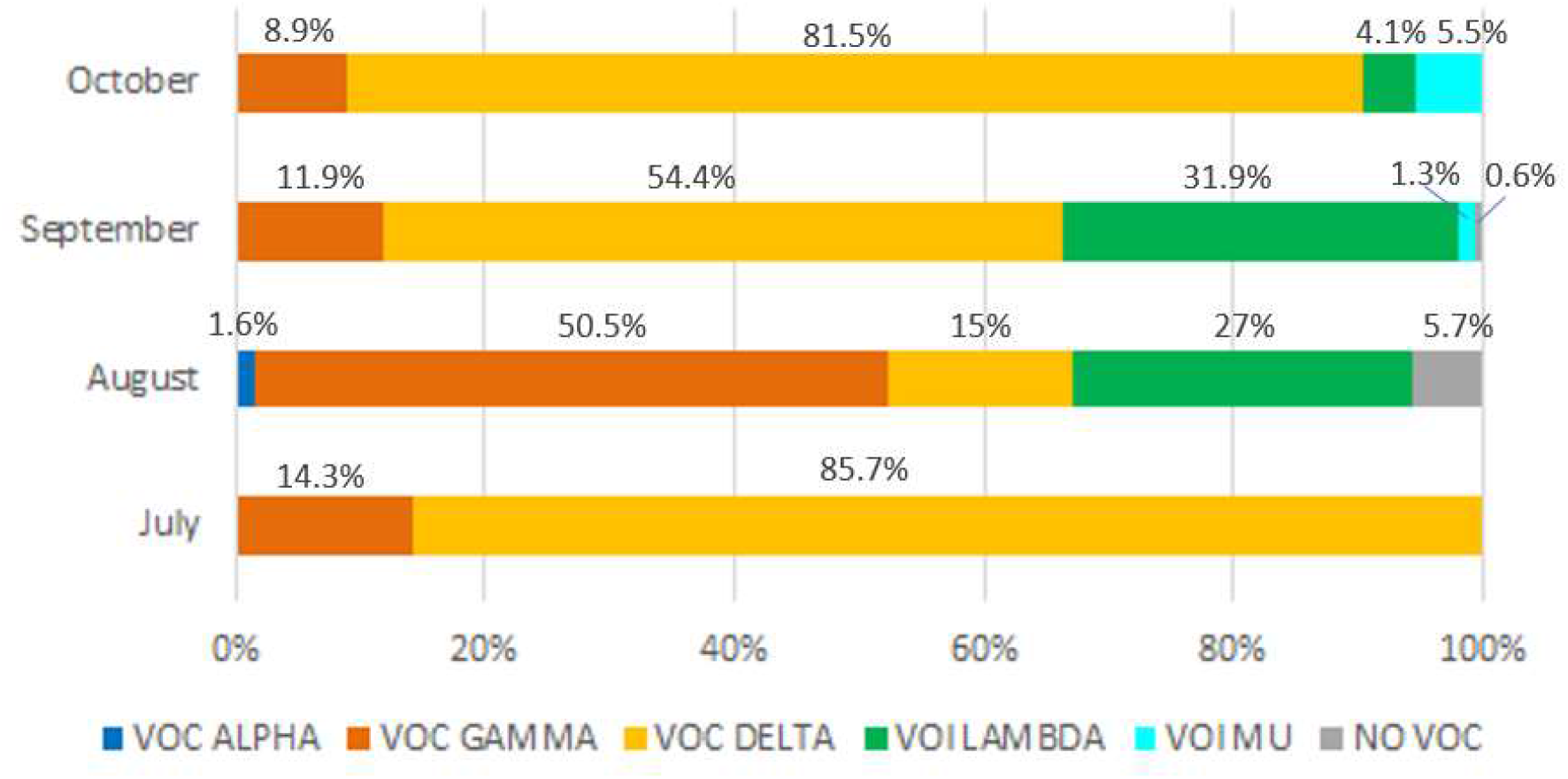
Distribution of SARS-CoV-2 variants of concern (VOC) and interest (VOI) in travellers, their close contacts and the study of Delta outbreaks in Córdoba Province, Argentina, by real time RT-PCR, July to October 2021.

## Discussion

The VOC/VOI detection strategy using consecutive real-time RT-PCRs for detection of relevant mutations implemented in our laboratory resulted a very useful and cost-effective diagnostic tool for the typing of variants. It was possible to monitor the variation of VOC/VOI distribution in the population over time, as well as to perform surveillance in travellers, which allowed an early detection of VOC Delta, making it possible to take measures to prevent its spread.

In Argentina, variant surveillance of SARS-CoV-2 and the study of variant spread is performed by national entities (National Ministry of Health and National Ministry of Science and Technology), using WGS or partial sequencing of the Spike protein, in a limited number of centres equipped with the necessary infrastructure. For this, positive traveller’s samples and randomly selected positive samples from each province are sent for processing to the laboratory in charge of carrying out the sequencing. The time between sending the samples and obtaining the result is between 2 and 15 days. Although WGS and partial sequencing provide accurate information (mainly WGS), they are laborious, time consuming, expensive and require extensive data processing, which has led to the search for faster and simpler alternatives for VOC/VOI detection [Ong et al. 2021, Lind et al. 2021]. In this sense, the strategy implemented during this work allowed to process a high number of samples in a short period of time. While the number of complete genomes from Córdoba province reached 140 during the studied period [Proyecto PAIS, Ministerio de Salud de la Nación Argentina 2021], the samples processed by the real time RT-PCR for detection of VOC/VOI was 3941. In addition, sample processing was considerably faster, without the need of sample derivation to the sequencing center; furthermore, costs were lower. In this way, Córdoba is the first province to implement this strategy for screening of VOC/VOI in Argentina, providing a quick and simple methodology that can be transferred to other laboratories that require it.

Using these real time RT-PCRs, it was possible to monitor variant circulation in the community since the introduction of VOC Alpha and Gamma in February and March 2021, respectively. Thus, in the following months, it was observed that, unlike the USA and Europe, VOC Gamma had an exponential increase and gained prominence over VOC Alpha and other variants, as happened in neighbouring countries, such as Brazil and Chile [Brazil Mutation Report and Chile Mutation Report]. Moreover, results obtained showed concordance with those obtained by WGS [Ministerio de Salud de la Nación Argentina 2021], showing the usefulness of the strategy developed to monitor frequency and variations of VOC/VOIs.

On the other hand, this strategy was also a very useful tool for the surveillance of SARS-CoV-2 variants in travellers, being able to detect VOC quickly and easily, and enabling rapid detection of Delta variant in Córdoba province. Rapid identification of VOC is key to taking measures to prevent their spread. In this case, the early detection of Delta allowed to isolate case zero and all its close contacts, circumscribing the outbreak and delaying the virus spread in our province. Subsequently, Delta began to be detected in the community, gradually increasing its proportion throughout the months studied, which differed from what happened in other parts of the country, where an abrupt increase was registered (frequency of VOC Delta increased in a short period of time) [9]. This coincided with the decrease in the number of SARS-CoV-2 positive cases [14], unlike what was observed in other parts of the world [4, 7], where the sustained advance of the VOC Delta drove new waves of infections [15]. Some reasons that could explain this difference include the exhaustive case identification, study and isolation of close contacts of positive Delta cases carried out by the Government of the Province, different epidemiological scenarios (Delta entered into our region with a particular variant circulation, in which VOC Gamma presented the highest frequency), acquired immunity of the population associated with the second wave (occurred on May-July 2021 in our province), and different vaccination programs implemented in the countries.

Former VOIs Zeta and Epsilon were detected in low percentages in the period studied, with currently no detection, supporting the new WHO classification, in which they are no longer designated as variants of interest or variants under surveillance [WHO 2021]. VOI Lambda was the second major variant of circulation in the community, which gradually increased its circulation, coinciding with what was reported at the national and regional levels [Outbreak info 2021, PAIS 2021, Ministerio de Salud de la Nacion 2021]. In the last month studied (October) this variant decreased its frequency slightly, probably as a consequence of the increase of VOC Delta in the community. In the coming weeks we will be able to observe if VOC Delta definitely displaces the rest of the variants, as has happened in many parts of the world (Ourbreak info 2021), or if the balance in the frequency of variants is maintained, continuing with a particular local epidemiological scenario.

The strategy described here present some limitations: a)-some samples show inconclusive results, b)-samples with Ct values >30 cannot be typified, c)-many diagnostic PCR platforms can deplete swab material, leaving an inadequate volume of residual sample for a multitube mutation screen [Wang et al. 2021], d)-since VOC/VOI classification is dynamic and is constantly changing [WHO 2021], the strategy must be constantly reviewed and evaluated in order to corroborate whether the algorithm used is adequate for a correct VOC typing, e)-it is not possible to find mutations other than those specifically searched for, which leads to not being able to detect new emerging VOC/VOIs. These limitations show that WGS cannot be replaced by real time RT-PCR specific for VOC/VOI. Furthermore, these methodologies complement each other. While specific real time RT-PCRs for mutations of interest are a useful tool for rapid VOC/VOI screening, WGS-based parallel surveillance is critical to detect new emerging variants and to study phylogeographic relationships between circulating viruses.

In conclusion, we report a valid strategy based on real time RT-PCR for VOC/VOI detection, first implemented in Argentina, which balance cost and time processing, capable to monitor currently recognized variants of concern. In the present moment requiring rapid strain typing to guide public health measures, such a rapid and accessible approach is essential.

## Data Availability

All data produced in the present study are available upon reasonable request to the authors

## Ethical statement

Ethical review and approval were not required for the study on samples obtained from human participants in accordance with the local legislation and institutional requirements. Written informed consent from the participants legal guardian/next of kin was not required to participate in this study in accordance with the national legislation and the institutional requirements.

